# A Mendelian randomization study of insulin therapy for type 1 diabetes increasing the potential risk of ovarian cancer

**DOI:** 10.1101/2024.07.19.24310599

**Authors:** Xue Bai, Ling Zhang

**Affiliations:** Ji Lin University

**Keywords:** Type 1 diabetes (T1D), Insulin product, Ovarian cancer (OC), Mendelian randomization (MR)

## Abstract

**Background:** Type 1 diabetes (T1D) has been associated with a higher risk of Ovarian cancer (OC), albeit the mechanisms underlying this association remain elusive. A better understanding of the relationship between T1D and OC may contribute to improved primary prevention of OC. We aimed to investigate the putative causal role of T1D on OC, and to identify the potentially mediatory effects of the usage of insulin product underlying this relationship.

**Methods:** We performed a two-sample Mendelian randomization (MR) analysis using genetic variants associated with T1D and OC from genome-wide association studies. Then, a multivariable MR analysis was conducted to investigate whether T1DM has an independent effect on OC after adjusting for potential confounders. Finally, the mediating role of insulin product was subsequently explored using mediation analysis via two-step MR.

**Results:** the MR estimated based on IVW method indicated a causal association between genetically determined T1D and Ovarian cancer (OC) (OR: 1.0006, 95% CI 1.0001–1.0011; *P* = 0.0164). After adjusting for body mass index, Smoking, physical activity, age at menopause and age at menarche, respectively, we found that a causal relationship between T1DM and OC was still statistically significant (OR>1, *P* <0.05). The two-step MR analysis revealed that insulin product acted as a mediating moderator between the T1D and OC (mediated proportion, 1.07%).

**Conclusions:** Our findings suggest that T1D may confer a risk effect to OC, mediated in part by therapeutic insulin product. Therefore, precise dosage of insulin product or an alternative to insulin in T1D patients have a profound significance in terms of the prevention of OC.

## 1. Introduction

Ovarian cancer (OC) is defined as an extremely threatening gynecological malignancy with crucial global impact on women’s well-being and economic burden. It stands as a dominating cause of female mortality worldwide, with a miserable escalation in mortality rates[1]. The absence of early clinical feature and the dearth of sensitive and specific diagnostic indicators system lead to the diagnosis of advanced-stage disease in over 70% of ovarian cancer patients[2]. Furthermore, it is worth notice that the 5-year survival rate for individuals diagnosed with advanced-stage ovarian cancer stands at approximately 25%, an extraordinarily lower figure compared to the 5-year survival rate of individuals with early-stage ovarian cancer, which reaches 92% [3]. Despite the advancement of screening methods and treatment modalities, survival rates for patients with Ovarian cancer have not ameliorated. Consequently, exploring the etiology and higher risk factors of ovarian cancer are of great significance for prevention and treatment of disease actively and effectively.

The pathogenesis of ovarian cancer is still a blue ocean of medical research. Reported nosogenesis include genetic factors, endocrine factors, fertility factors, immune factors, chronic inflammation, oxidative stress, and environmental factors[4][5][6]. Type 1 diabetes (T1D) is an autoimmune and metabolic disease that results in pancreatic β-cell damage, giving rise to a decrease in insulin synthesis or a complete lack of insulin secretion in patients, which contributes to elevated blood glucose[7]. Over the years, countless scientists have been devoting themselves to explore the relationship between Type 1 diabetes and ovarian cancer. There is sufficient biological evidence that overexpression of IGF-I is demonstrated in ovarian cancer[8][9]. Besides, low serum sex hormone-binding globulin[10][11], inflammatory pathways[12][13], leptin overproduction[14][15],and microRNAs[16][17][18][19][20],may explain part of the mechanisms associating Type 1 Diabetes to Ovarian cancer. Epidemiologic evidence also suggests that people with Type 1 diabetes are at significantly higher risk for many forms of cancer[21][22][23][24][25]. Although there have been basic experimental researches and epidemiological investigation [8–24], regarding the association between Type 1 diabetes and ovarian cancer, there are no randomized clinical trials. Meanwhile, many studies have a great deal of confounding factors, with a high degree of heterogeneity and a deficiency of causality inference. However, due to the profound impact of Ovarian cancer on women’s health and quality of life, it is important to explore the causal association between Type 1 diabetes and ovarian cancer. When a causal relationship between Type 1 diabetes and ovarian cancer is established, it will provide a new clinical strategy for the management of ovarian cancer.

Traditional observational studies may cause deflection and even misjudgment of the research results due to various observable and unobservable confounding factors, reverse causality and bias; meanwhile, they principally focus on the correlation between exposure factors and outcomes instead of the actual causality. Mendelian randomization (MR) is a new epidemiological method that imitates the design of randomized controlled studies[26]. It uses single nucleotide polymorphisms (SNPs) as instrumental variables (IVs) to infer causal relationship between the risk factors and interested outcomes. SNP is randomly assigned to individuals with gametes during meiosis [27]. which is similar to the essential demands of randomized controlled trials. Meanwhile, genovariation precedes the occurrence of diseases, which avoids the potential influence of reverse causality. Consequently, MR is an ideal way to explore the causal relationship between Type 1 diabetes and Ovarian cancer.

This study used a two-sample MR and Mediated Mendelian randomization analyses design to investigate whether Type 1 diabetes has a causal association with Ovarian cancer and estimate the mediating roles between them to provide scientific evidence for Ovarian cancer prevention and treatment.

## 2. Materials and methods

### 2.1 Experimental design

The current investigation selected Type 1 diabetes (T1D) as the exposure factor and extracted instrumental variables (IVs) from the exposure dataset in the shape of single nucleotide polymorphisms (SNPs) that revealed conspicuous associations with T1D. The outcome variable of interest was ovarian cancer (OC). MR analysis was conducted to investigate the causal relationship between the exposure and outcome variables. To evaluate heterogeneity and dispose potential issues of horizontal pleiotropy, Cochran’s Q test, MR Presso’s test and MR Egger’s test were preferred. Furthermore, sensitivity analysis was implemented to ascertain the robustness and reliability of the expected results [28]. The reliability of MR analysis is predicated on the implementation of three conditional assumptions [29]: (i) There exists a firm correlation between instrumental variables and exposure factors. (ii) No confounding factors are existent that might affect the relationship between exposure and outcome variables, especially, no genetic polymorphisms. (iii)Instrumental variables do not exert a direct influence on the outcome, but exclusively effect the outcome via the exposure factors. Refer to Figs 1 for a visual depiction of this assumptions.

**Fig. 1.**
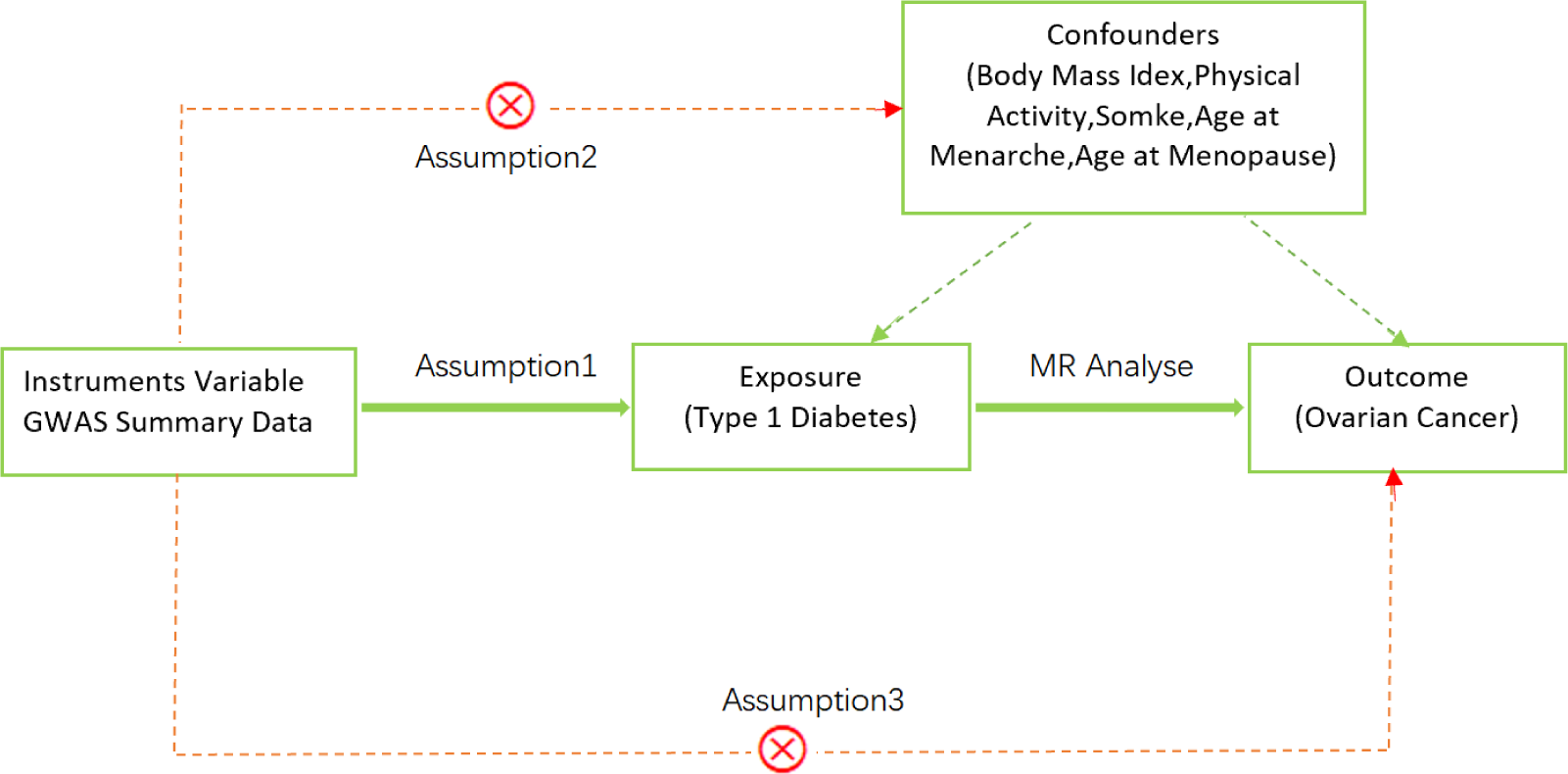
A diagram of our main MR study. Assumption1, there exists a firm correlation between instrumental variables and exposure factors; Assumption2, No confounding factors are existent; Assumption3, Instrumental variables exclusively effect the outcome via the exposure factors.

### 2.2 Data sources

All data were independently acquired from the IEU GWAS database (https://gwas.mrcieu.ac.uk/terms/), founded by the University of Bristol, UK. Detailed information is provided in Table 1.

**Table. 1.** An overview of the GWAS datasets used in the MR analysis.

| Contribution | Traits | Sample size | Number of SNPs | Author | Population | GWAS ID |
| --- | --- | --- | --- | --- | --- | --- |
| Exposure | Type 1 diabetes | 457,695 | 24,182,422 | Sakaue | European | ebi-a-GCST90018925 |
| Confounders | Body mass index | 461,460 | 9,851,867 | Ben Elsworth | European | ukb-b-19953 |
|  | smoked | 461,066 | 9,851,867 | Ben Elsworth | European | ukb-b-20261 |
|  | Age at menopause | 143,819 | 9,851,867 | Ben Elsworth | European | ukb-b-17422 |
|  | Age at menarche | 182,416 | 2,441,816 | Perry JR | European | ieu-a-1095 |
|  | physical activity | 377,234 | 11,808,007 | Klimentidis | European | ebi-a-GCST006097 |
| Mediators | insulin product | 462,933 | 9,851,867 | Ben Elsworth | European | ukb-b-15445 |
| Outcome | Ovarian cancer | 199,741 | 9,822,229 | Burrows | European | ieu-b-4963 |

### 2.3 Instrumental variables selection

Initially, SNPs revealing significant associations with exposures (selection criteria: P<5.0×10−8) were selected based on the 1,000 whole-genome European population. The linkage disequilibrium parameter (r2) was set at 0.001, and the genetic distance was founded in 10,000 kb to insure the independence of each instrumental variable (IV), excluding interference from other IVs. Subsequently, SNPs with strong linkage disequilibrium (r2 > 0.8) were applied to compensate for missing SNPs in the outcome dataset. Finally, exposure and outcome data were combined while keeping the corresponding values for exposure and outcome with the same effect alleles[30]. The residual SNPs constitute the ultimate instrumental variables connected with the exposure. To consider the proportion of SNP phenotypic variation in the Mendelian randomization process, we calculated R2 values via using the formula R2 = 2 ×β2 × EAF ×(1-EAF)/[2 × β2 × EAF × (1-EAF)+ SE2 × 2 × N × EAF × (1-EAF)], where β is the effect estimate of the genetic variant in the exposure GWAS, EAF represents the Allele 1 frequency, SE represents the standard error, and N represents the sample size[31][32]. And then, the instrumental strength of SNPs for each socioeconomic trait was calculated using the F-statistic, evaluated as F =[(N—k—1)/k] × [R2/ (1—R2)], where N is the sample size, k represents the total number of SNPs, and R2 represents the total proportion of phenotypic variation interpreted by all the SNPs in the MR analysis. An F-statistic greater than 10 manifests that the combined SNPs serve as sufficiently strong instruments to illuminate phenotypic variation, while an F-statistic of 10 or less elucidates a weak instrument[32].

### 2.4 Mendelian randomization

MR analysis is a advantageous tool in epidemiological studies. The study began with a two-sample Mendelian randomization employing T1D and confounders (including body mass index, Somke, physical activity, age at menopause, age at menarche) as exposures and OC as an outcome indicator severally. As T1D and confounders are correlated in clinical studies, multivariate Mendelian randomization was performed to correct for the results. Inverse variance weighted (IVW), weighted median estimator (WME) and MR-Egger regression were the main methods in Mendelian randomization.

### 2.5 Mediated Mendelian randomization

To explore the role of insulin product in the relationship between T1D and OC, we used a two-step MR approach, as shown in Fig. 2. The two-step approach is considered less tendency to the biases inherent in common multivariate methods [33].In MVMR, the total effect of each exposure is accounted for by direct effect and an indirect effect. There is a mediator if the following conditions were met:1) there was a correlation between T1D and the mediator (β 1); 2) the mediator was correlated with OC (β 2). 3) T1D was closely related to OC but not adjusted for the mediator (β3’); The mediation ratio was calculated as (β 1 x β 2)/(β 3’). β 1 x β 2 is the indirect effect and the total effect(β3) is β 3’ + β 1 x β2.

**Fig. 2.**
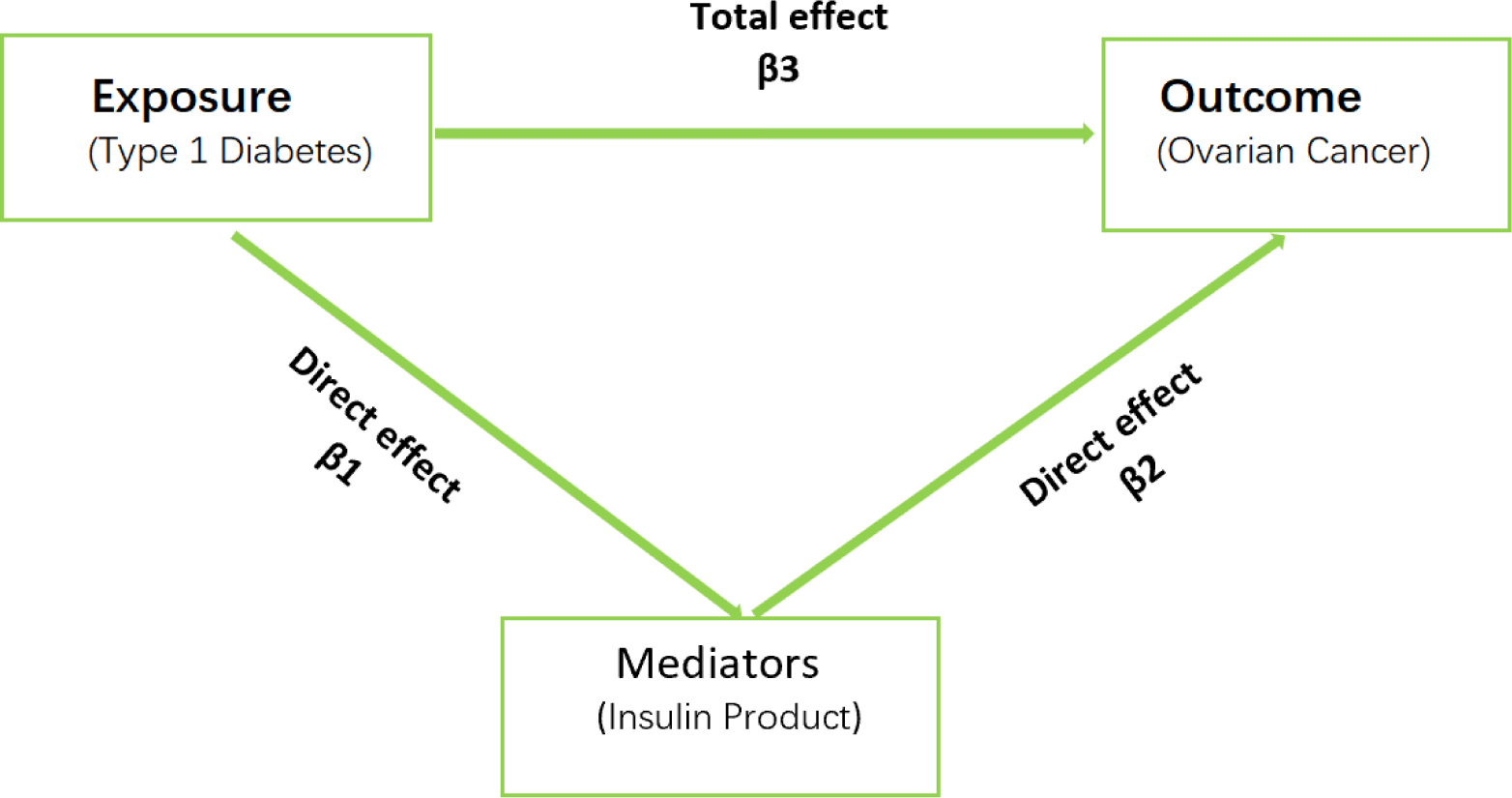
A diagram of mediator MR analysis. This diagram illustrates the potential casual pathway from T1D to OC, with Insulin Product serving as mediators.

### 2.6 Sensitivity analysis

This study implemented a ‘leave-one-out’ sensitivity analysis, whereby individual SNPs were removed one at a time, to assess whether the observed variation was dominant the association between the exposure and outcome variables. Secondly, to confirm the presence of horizontal pleiotropy in this MR analysis, the MR-Egger intercept test was absorbed. If the intercept term in the MR-Egger intercept analysis did not prove statistically significant, there is no horizontal pleiotropy within the study [34].Finally, Cochran’s Q statistic was implemented in this study to ascertain heterogeneity. A significant level of heterogeneity within the analysis.was manifested by a statistically significant result from the Cochran’s Q statistic test. However, when the number of SNPS is greater than 3, IVW may have certain heterogeneity tolerate.

### 2.7 Statistical analysis

In this study, the “TwoSampleMR” and “Mendelian randomization” [35].Packages accessible within RStudio software were used to carry out the exposure and outcome analyses. The outcomes from the Mendelian randomization (MR) analysis were defined as beta (β) values, indicating the impact of T1D and insulin product on OC. Furthermore, the associated 95% confidence intervals (CI) were recorded for all causal estimates. A significance threshold of p < 0.05 was applied to discern statistical significance.

## 3 Results

### 3.1 Two-sample Mendelian randomization

In the IVW models, compelling evidence indicated a significant association between Type 1 Diabetes and the risk of ovarian cancer (OR: 1.0006, 95% CI 1.0001–1.0011; *P* = 0.0164). Additionally, we assessed the association of the 17-SNP instrument with OC using MR-Egger, weighted median. MR-Egger regression showed an OR of 1.0009 (95% CI 1.0001– 1.0011; *P* = 0.0492), while weighted median (OR 1.0009, 95% CI 1.0002– 1.0016; *P* = 0.0087) techniques provided directionally consistent results. These findings suggest that the results remain robust even in the face of potential violations of MR assumptions. Furthermore, no evidence of pleiotropy or heterogeneity was detected in the MR-PRESSO global test, MR-Egger intercept test, and Cochran’s Q test. The data is summarized in Supplementary Tables S1-2. To obtain MR estimates for each individual SNP, we conducted the analysis multiple times for each exposure-outcome combination. In each iteration, a different single SNP was used for the analysis. The scatterplots depicted in Fig.3 illustrate that SNPs exhibiting a larger effect on Type 1 Diabetes also exert a greater impact on the risk of ovarian cancer. Each method is expressed by a different colored line, with the slope of the line manifesting the estimated causal effect. A leave-one-out analysis is feasible, wherein the MR analysis is repeated while excluding each SNP independently. This allows us to explicit that a majority of the associated signals were not primarily impacted by a single genetic marker, as is demonstrated in Fig.4.

**Fig 3.**
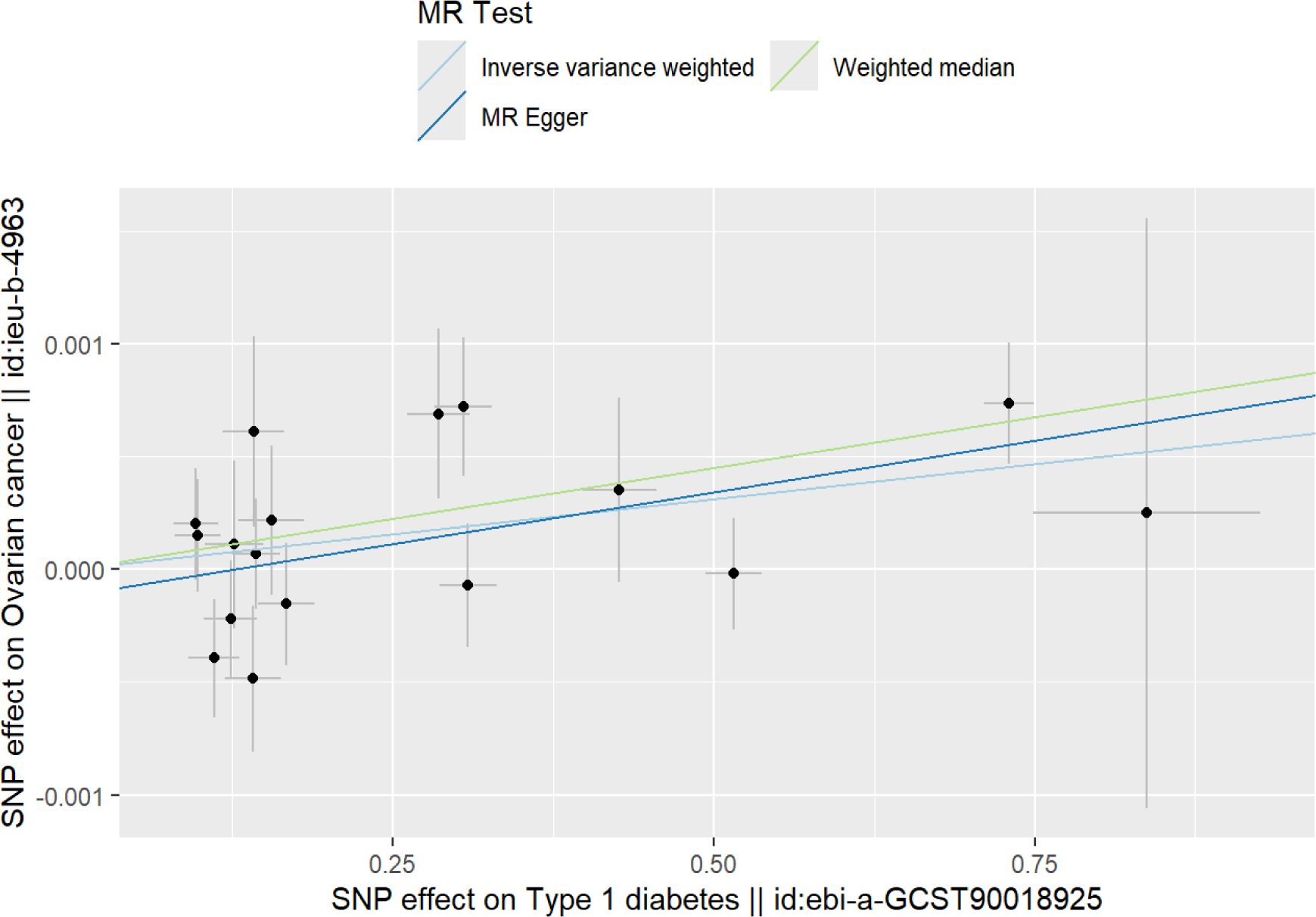
Scatter plot of single SNP withT1D as the exposure and OC as the outcome.

**Fig 4.**
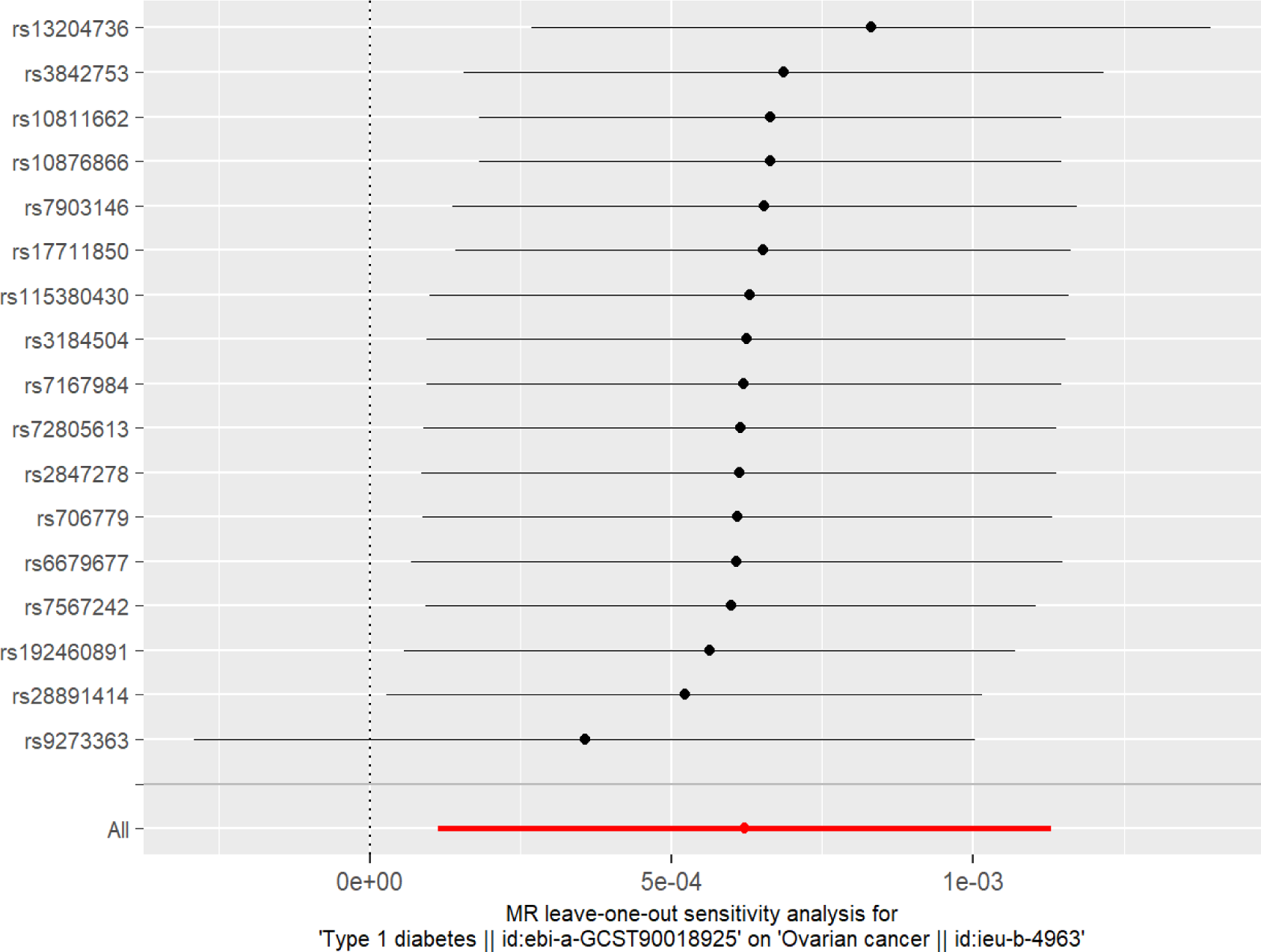
Leave-one-out analysis of single SNP withT1D as the exposure and OC as the outcome.

**Table. 2.**
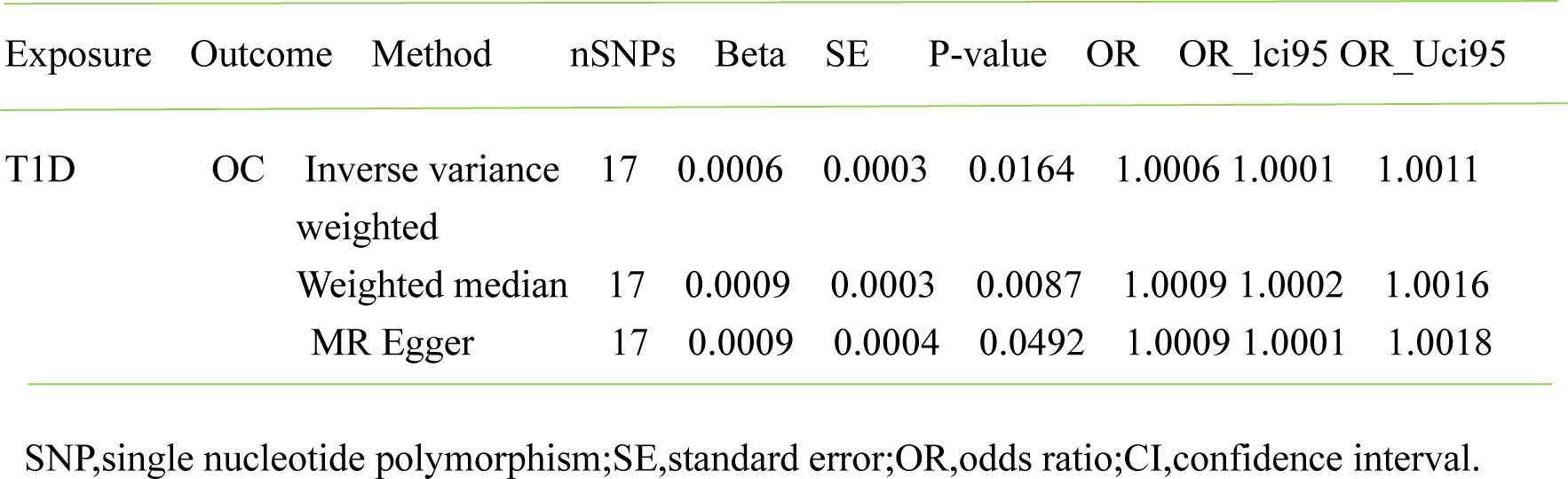
Two-sample MR analysis results under different methods.

### 3.2 Multivariate Mendelian randomization

To adjust for potential pleiotropic pathways that might confound the relationship between T1D and OC, an MVMR model was utilized. In this model, the combined effect of T1D on body mass index, Smoking, physical activity, age at menopause and age at menarche respectively was used as an exposure for OC outcomes. The results of the IVW analysis revealed that the previously significant correlation between T1D and OC was not attenuated in the MVMR model and was still statistically significant, detailed results are presented in Table 3. Furthermore, There is no pleiotropy or heterogeneity in the MR-PRESSO global test, MR-Egger intercept test, and Cochran’s Q test (Supplementary Tables S1-2).

**Table. 3.**
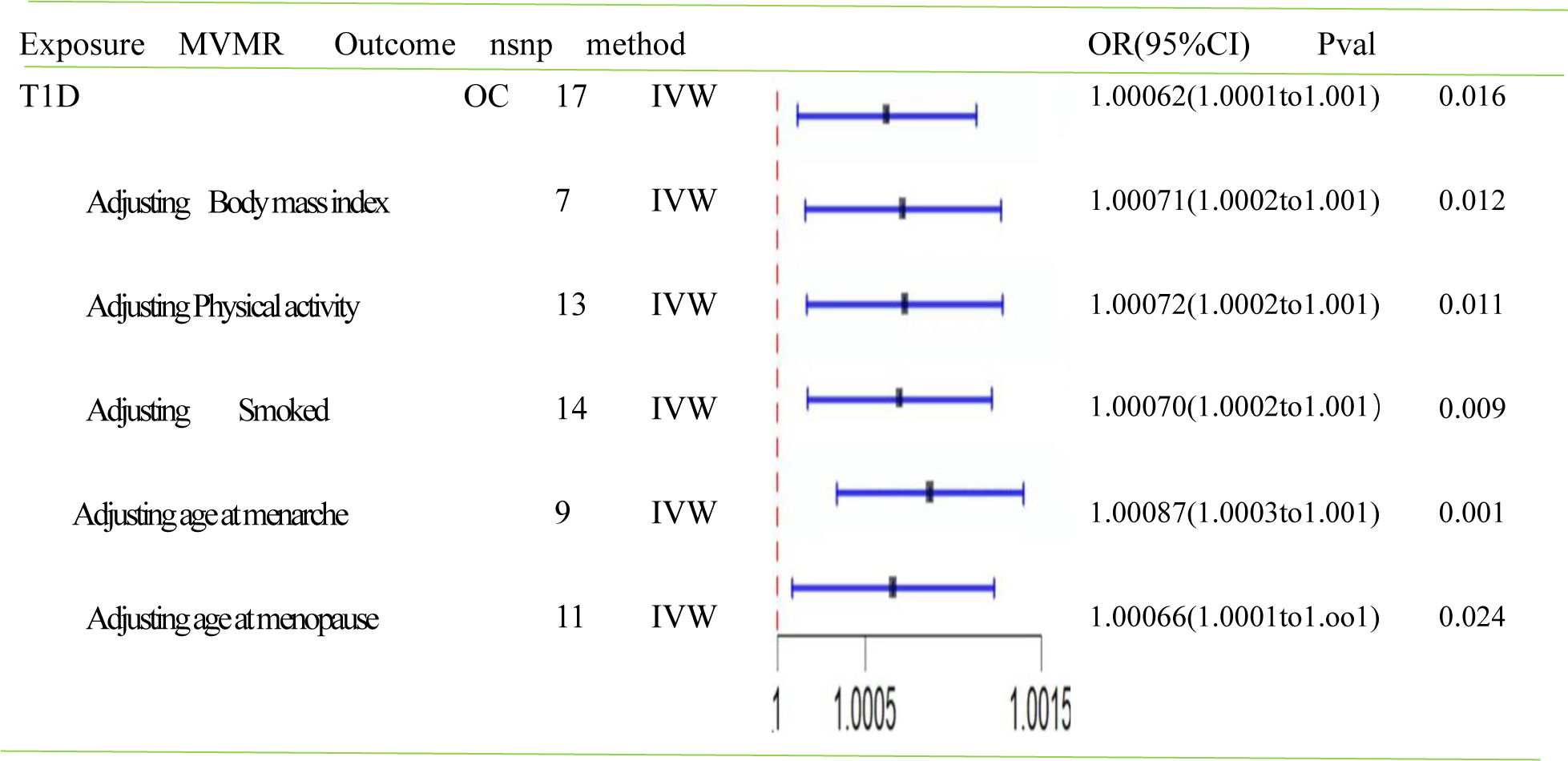
Forest plot of the MVMR analysis. The direct causal effect of T1D on OC risk was determined by adjusting for traits such as Body mass index, Physical activity, Smoked, Age at menarche, Age at menopause.

### 3.3 Mediated Mendelian randomization

In order to investigate the mediating role of insulin product in the relationship between T1D and OC, a mediated MR analysis was conducted. The results of this study are detailed in Table 4. Our study revealed that insulin product serves as a mediator in the relationship between T1D and OC, with the mediating effect estimated to be (β = 6.63×10-4, 95% CI 2.10×10-4 to 1.12×10-3,*P* = 0.04).

**Table. 4.**
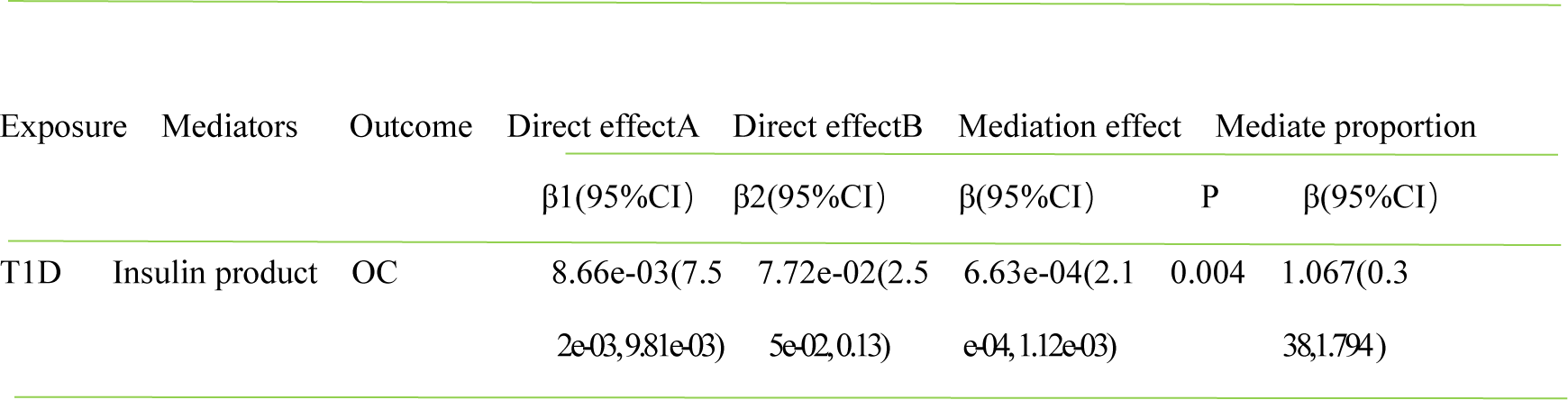
The mediation effect of mediators on the causal effect of T1D and OC.

## 4. Discussion

Ovarian cancer and Type 1 diabetes bring tremendous security challenges to public health. The relationship between T1D and OC remains a subject of debate, with the mechanism of this association not fully revealed. In this study, we utilized GWAS database and employed two-sample MR to examine the causal relationship between T1D and OC. The findings indicate that T1D may potentially accelerate the progression of OC. The interconnectedness between T1D and OC was mediated by insulin product. Insulin product emerges as a risk factor for OC, underlining the precise control of insulin medicine to mitigate the risk of OC.

In recent decades, several observational studies have sought the link between T1D and OC. Nonetheless, since there is inherent nature of recall and select bias in observational study, certain biases might contribute to inaccurate reporting of causal relationship[36]. Several cohorts [37][38][39][40]and case–control [41]studies have been reported that a history of T1D is associated with an augmented risk of OC, however, other relevant studies found a negative significant association [42][43][44][45]. Besides, a subgroup meta-analysis based on DM types indicated that the risk of OC in T1DM group (44%) is higher than in T2DM group (17%) [46]. However, MR analysis overcomes the limitations of observational studies, which unambiguously interpret the causal relationship between T1D and OC. Moreover, the positive link was even more prominent in studies that adjusted for confounding factors (such as, body mass index, smoking, physical activity, age at menopause and age at menarche) than these for unadjusted analysis.

The underlying carcinogenesis effect of T1D to OC was not completely revealed at present, but several plausible mechanisms have been used to explain the links between them. Previous studies have shown that the neoplastic process has been considered to influenced by T1D through these mechanisms, mainly including hyperglycemia, adipokine imbalances, and chronic inflammation[47][48][49]. Because of a prolonged exposure to inflammation and hyperglycemic condition, the repair cycles which are associated with incessant ovulation process could be slowed down, thus, resulting in an underlying risk of OC[50]. In addition, hyperglycemia may lead to endothelial dysfunction, endothelial cell death and aberrant neoangiogenesis, ultimately, contributing to cancer progression[51][52]. Studies have shown that adipose metabolic dysregulation as an hallmark of T1D, can result in high levels of inflammatory cytokines, including IL-6 and TNF-α [49] These cytokines stimulate cell proliferation, invasion, and evasion of antitumor immunity pathways [13].

Mediated Mendelian randomization was employed to delve into the potential mechanism underlying this relationship. We find that therapeutic insulin product as a mediator could explain parts of this relationship. This is consistent with the results of some observational studies. Since 1972 in the animal experimental researches[53][54], it has been recognized that the insulin has promoted cancer growth due to the mitogenic effect of insulin by alteration in the PI3K/AKT pathway and mitotic kinase pathway [55]. Type 1 diabetes patients have widely used insulin replacement therapy to strictly control the metabolic status and prevent the progression of long-term complications associated with persistent hyperglycemia [56]. Although there are structural similarities between exogenous and endogenous insulin, the insulin replacement therapy is non-physiologic, and thus, a supraphysiologic dose of insulin needs to be injected in order to achieve the Physiological needs. This contributes to an inevitable therapeutic increase in the dose of insulin therapy, leading to exogenous systemic hyperinsulinemia [57]. Hyperinsulinemia increases the levels of estradiol and testosterone, which may result in cancer initiation and progression in patients [58][59].

The characteristic of our study is consistent with all MR assumptions. We integrated data from GWAS of both Type 1 diabetes and ovarian cancer. This robust methodology, coupled with extensive data sources, supports our causal inference by alleviating biases stemming from confounding and reverse causation. Furthermore, our findings provide a reliable theoretical framework for future investigations focused on improving therapeutic approaches for Type 1 diabetes, such as insulin preparation replacement reagent. This potential improvement may reduce the risk of ovarian cancer, carrying an important directive function for clinical management. However, there are several limitations in our research. On one hand, this investigation is the lack of comprehensive GWAS in non-European ancestries. Therefore, the assessment of these causal pathways in diverse ethnic groups should be paid to in the future studies. On the other hand, the utilization of pooled data from genome-wide association studies was short of individual-specific information, eliminating subgroup analyses for variables such as age, disease duration, treatment method, disease classification, and son on. This limitation makes it impossible to compare potential differences in causal effects among these subgroups.

## 5. Conclusion

In conclusion, our study provides evidence supporting a causal relationship between T1D and OC. Furthermore, therapeutic insulin product mediated the effect between T1D and OC. Therefore, precise dosage of insulin product or an alternative to insulin in T1D patients have a profound significance in terms of the prevention of OC.

## Data Availability

All data produced in the present study are available upon reasonable request to the authors
All data produced in the present work are contained in the manuscript
All data produced are available online atGWAS summary data.

## Notes

### Competing Interest Statement

The authors have declared no competing interest.

### Funding Statement

This study research was funded by The National Natural Science Foundation of China (Grant No. 82273479 to LZ)

### Author Declarations

JiLin University Ethics Committees

