## supplementary Tables S1-2 for "A Mendelian randomization study of insulin therapy for type 1 diabetes increasing the potential risk of ovarian cancer"

### Supplementary TablesS1

F-stat for all Exposures

| Contribution | Traits | nIV | R2 | F_stat |
|---|---|---|---|---|
| Exposure | Type 1 diabetes | 18 | 0.006994119 | 179.09 |
| Confounders | Body mass index | 458 | 0.06321212 | 67.92 |
| | physical activity | 19 | 0.001732107 | 34.448 |
| | smoked | 84 | 0.007239536 | 40.019 |
| | Age at menarche | 68 | 0.03107539 | 86.004 |
| | Age at menopause | 114 | 0.06881494 | 93.156 |
| Mediators | insulin product | 9 | 0.004501733 | 232.6 |
| | metformin | 46 | 0.005862344 | 59.339 |
| | IGF-1 | 349 | 0.09859128 | 107.21 |
| | Fasting blood glucose | 22 | 0.03505103 | 95.848 |
| | Blood glucose | 114 | 0.03918007 | 143.2 |
| | Total cholesterol | 61 | 0.0764044 | 155.98 |
| | LDL cholesterol | 48 | 0.08796713 | 231.15 |
| | HDL cholesterol | 346 | 0.1948019 | 279.97 |
| | Triglyceride | 293 | 0.1267692 | 216.64 |
| | Total testosterone | 215 | 0.07720643 | 89.596 |
| | Bioavailable testosteror | 96 | 0.0509005 | 99.822 |
| | Sex hormone-binding g | 376 | 0.1793203 | 230.51 |
| | Estradiol | 13 | 0.006507103 | 74.403 |
| | leptin | 5 | 0.005928548 | 67.633 |
| | C-reactive protein | 9 | 0.01307593 | 122.21 |
| | Lymphocyte count | 519 | 0.1133434 | 129.16 |

### Supplementary TablesS2

| exposure | confounders | outcome | method | nsnp | b | se | pval | lo_ci | up_ci | or | or_lci95 | or_uci95 | Q | Q_df | Q_pval | egger_intercept | egger_intercept_ | egger_intercept_ | presso_global_ |
|---|---|---|---|---|---|---|---|---|---|---|---|---|---|---|---|---|---|---|---|
| Type 1 diabetes | | Ovarian cancer | Inverse variance weighted | 17 | 0.000621589 | 0.0002591 | 0.01643862 | 1.14E-04 | 0.00112943 | 1.000622 | 1.000114 | 1.00113 | 19.46914 | 16 | 0.2450868 | -0.000114268 | 0.0001317 | 0.3991264 | 0.281 |
| | | | Weighted median | 17 | 0.000900215 | 0.000343071 | 0.00869059 | 2.28E-04 | 0.00157264 | 1.000901 | 1.000228 | 1.001574 | | | | | | | |
| | | | MR Egger | 17 | 0.000915882 | 0.00042798 | 0.04919663 | 7.70E-05 | 0.00175472 | 1.000916 | 1.000077 | 1.001756 | | | | | | | |
| Body mass index | | Ovarian cancer | Inverse variance weighted | 432 | 0.001803016 | 0.00078316 | 0.02132281 | 0.00026801 | 0.00333802 | 1.001805 | 1.000268 | 1.003344 | 465.0714 | 431 | 0.1242917 | 7.18E-06 | 3.85E-05 | 0.8521136 | 0.1345 |
| | | | Weighted median | 432 | 0.001476838 | 0.00137674 | 0.28340353 | -0.0012216 | 0.00417525 | 1.001478 | 0.9987792 | 1.004184 | | | | | | | |
| | | | MR Egger | 432 | 0.001434525 | 0.00212536 | 0.5000668 | -0.0027312 | 0.00560023 | 1.001436 | 0.9972725 | 1.005616 | | | | | | | |
| physical activity | | Ovarian cancer | Inverse variance weighted | 19 | -0.00346012 | 0.00461145 | 0.4530551 | -0.0124986 | 0.00557833 | 0.9965459 | 0.9875792 | 1.005594 | 19.97437 | 18 | 0.3342645 | 0.000318754 | 0.0003231 | 0.3376276 | 0.3335 |
| | | | Weighted median | 19 | 0.000485547 | 0.00593283 | 0.9347734 | -0.0111428 | 0.0121139 | 1.0004857 | 0.988919 | 1.012188 | | | | | | | |
| | | | MR Egger | 19 | -0.02448334 | 0.02180071 | 0.2770259 | -0.0672127 | 0.01824605 | 0.9758139 | 0.9349963 | 1.018414 | | | | | | | |
| smoked | | Ovarian cancer | Inverse variance weighted | 78 | 1.12E-05 | 0.00440835 | 0.9979775 | -0.0086292 | 0.00865154 | 1.0000112 | 0.9914079 | 1.008689 | 65.38105 | 77 | 0.8247321 | 4.88E-05 | 0.0001464 | 0.7395161 | 0.822 |
| | | | Weighted median | 78 | 2.34E-03 | 0.00654657 | 0.7201991 | -0.0104864 | 0.01517622 | 1.0023477 | 0.9895684 | 1.015292 | | | | | | | |
| | | | MR Egger | 78 | -6.89E-03 | 0.02114649 | 0.7454323 | -0.0483377 | 0.03455651 | 0.9931331 | 0.9528119 | 1.035161 | | | | | | | |
| Age at menarche | | Ovarian cancer | Inverse variance weighted | 60 | -0.00045428 | 0.00078217 | 0.561376 | -0.0019873 | 0.00107877 | 0.9995458 | 0.9980146 | 1.001079 | 46.64192 | 59 | 0.8781744 | -5.94E-05 | 0.0001374 | 0.6670837 | 0.858 |
| | | | Weighted median | 60 | 0.000930334 | 0.00116002 | 0.4225523 | -0.0013433 | 0.00320397 | 1.0009308 | 0.9986576 | 1.003209 | | | | | | | |
| | | | MR Egger | 60 | 0.00078531 | 0.00297182 | 0.7925225 | -0.0050395 | 0.00661008 | 1.0007856 | 0.9949732 | 1.006632 | | | | | | | |
| Age at menopause | | Ovarian cancer | Inverse variance weighted | 105 | 0.001664489 | 0.00068749 | 0.0154733 | 0.00031701 | 0.00301197 | 1.001666 | 1.0003171 | 1.003017 | 104.0715 | 104 | 0.4795836 | 5.25E-05 | 5.96E-05 | 0.3801208 | 0.491 |
| | | | Weighted median | 105 | 0.001293125 | 0.00113508 | 0.2546051 | -0.0009316 | 0.00351788 | 1.001294 | 0.9990688 | 1.003524 | | | | | | | |
| | | | MR Egger | 105 | 0.000600921 | 0.00138908 | 0.6662069 | -0.0021217 | 0.00332351 | 1.000601 | 0.9978806 | 1.003329 | | | | | | | |
| Type 1 diabetes | | | | | | | | | | | | | | | | | | | |
| | Body mass index | Ovarian cancer | Inverse variance weighted | 7 | 0.000714221 | 0.00028449 | 0.01205547 | 0.0001566 | 0.0012718 | 1.000714 | 1.000157 | 1.001273 | 449.379641 | 6 | 0.0930529 | 2.01E-05 | 1.70E-05 | 0.2360119 | 0.104 |
| | physical activity | Ovarian cancer | Inverse variance weighted | 13 | 0.000725698 | 0.00028404 | 0.01062078 | 0.0001556 | 0.0012958 | 1.000726 | 1.000169 | 1.001283 | 27.655568 | 12 | 0.5363801 | 5.41E-05 | 0.000109 | 0.6201548 | 0.526 |
| | smoked | Ovarian cancer | Inverse variance weighted | 14 | 0.000695953 | 0.00026701 | 0.00914937 | 0.0001501 | 0.0012418 | 1.000696 | 1.000173 | 1.00122 | 79.050797 | 13 | 0.6892217 | -2.89E-05 | 3.22E-05 | 0.3697496 | 0.692 |
| | Age at menarche | Ovarian cancer | Inverse variance weighted | 9 | 0.000868718 | 0.0002711 | 0.00135311 | 0.0002798 | 0.0014576 | 1.000869 | 1.000337 | 1.001401 | 52.918137 | 8 | 0.8585937 | -5.01E-05 | 3.79E-05 | 0.1862377 | 0.887 |
| | Age at menopause | Ovarian cancer | Inverse variance weighted | 11 | 0.000658537 | 0.00029333 | 0.02476475 | 8.36E-05 | 0.0012335 | 1.000659 | 1.000084 | 1.001234 | 115.63368 | 10 | 0.3136846 | -6.07E-05 | 3.16E-05 | 0.055134 | 0.295 |
